# Antecedent flu-like illness and onset of idiopathic dilated cardiomyopathy: The DCM Precision Medicine Study

**DOI:** 10.1101/2024.10.04.24314926

**Authors:** Hanyu Ni, Jinwen Cao, Daniel D. Kinnamon, Elizabeth Jordan, Garrie Haas, Mark Hofmeyer, Evan Kransdorf, Jamie Diamond, Anjali Owens, Brian Lowes, Douglas Stoller, W. H. Wilson Tang, Mark Drazner, Palak Shah, Jane E. Wilcox, Stuart Katz, Javier Jimenez, Supriya Shore, Daniel P. Judge, Jonathan Mead, Jason Cowan, Patricia Parker, Gordon S. Huggins, Ray E. Hershberger

## Abstract

**Background:** Previous studies have speculated that a viral infection may act as a trigger in the development of idiopathic dilated cardiomyopathy (DCM) among individuals genetically at risk. This study aims to describe the frequency of DCM patients who reported experiencing symptoms of flu-like illness prior to their DCM diagnosis and to examine if this experience modified the association between genetics and DCM.

**Methods:** We analyzed data from the family-based cross-sectional DCM Study conducted between 2016 and 2021. Self-reported symptoms of flu-like illness proximal to DCM diagnosis were obtained from patient interview. Exome sequencing identified rare variants [pathogenic (P), likely pathogenic (LP), or uncertain significance (VUS)] in DCM genes. In a case-only design, logistic mixed models were used to examine if flu-like illness modified the effect of these rare variants on DCM risk. Firth logistic regression was used to examine if flu-like illness modified the effect of each of 13,400,141 common autosomal variants (minor allele frequency ≥1%) on DCM risk.

**Results:** Of 1,164 DCM patients, 30.2% reported symptoms of flu-like illness proximal to DCM diagnosis. The percentage of patients with antecedent flu-like illness by variant classification was 30.0% for P/LP, 29.6% for VUS only, and 30.0% for no P/LP/VUS. Antecedent flu-like illness was not found to modify the effect of carrying any P, LP or VUS variants on DCM risk (interaction relative risk =0.9, 95% CI: 0.7-1.3). However, significant modification of the effect of rs2102158 (3q24) by antecedent flu-like illness (p=2.74×10^−8^) was identified by case-only genome-wide association study (GWAS).

**Conclusions:** Approximately one-third of DCM patients experienced flu-like illness symptoms prior to DCM diagnosis. We did not find evidence that a flu-like illness modified the effect of rare variants on DCM risk; however, our GWAS analysis suggested that flu-like illness may modify the effect of a common variant on DCM risk.

**Clinical Perspective:**

1. What is new?

- This study is the first that provides a comprehensive epidemiologic profile of antecedent flu-like illness among DCM patients of diverse ancestry, recruited from geographically diverse heart failure programs across the United States.
- Antecedent flu-like illness was not found to modify the effect of harboring P/LP/VUS rare variants on DCM risk, even though approximately one-third of patients with DCM reported episodes of flu-like illness prior to their DCM diagnosis.
- GWAS analysis suggested that flu-like illness may modify the effect of a common variant at chromosome 3q24 on DCM risk.
2. What are the clinical implications?

- Results from this study will inform clinicians that no evidence was found to suggest that a flu-like illness modified the effect of harboring rare variants in DCM genes on DCM risk.
- A clinical presentation of DCM following a flu-like illness does not signal a higher or lower rare variant genetic risk for DCM and thus does not alter the need for recommended genetic testing.
- These findings do not address the long-held question of whether the clinical presentation of DCM can be caused by a flu-like illness in some patients; further investigation will be needed.

## INTRODUCTION

Idiopathic dilated cardiomyopathy (DCM) has been recognized as a complex disease resulting from many genetic determinants, possibly interacting with environmental factors.^1^ An unresolved question is whether a viral infection might cause the DCM or trigger its onset.^2^ Previous studies have linked DCM to genetic susceptibilities^1^ or viral infection,^3^ but most studies have examined genetics or viruses separately. It has been suggested that a viral infection may act as a trigger in the development of DCM, although the results have been inconclusive.^4–7^ Understanding the interplay of viral infection and genetic risk is essential for describing the etiology of DCM and in guiding its clinical treatment and prevention.

Influenza-like illness, also known as flu-like illness, is commonly identified by physicians based on patient reported flu-like symptoms and represents a medical diagnosis of possible influenza or other presumed viral illness causing a set of common symptoms. The World Health Organization defines an illness as a flu-like illness if the patient has a fever greater than or equal to 38 C° and a cough that began in the last 10 days.^8^ A case definition of fever of 100°F or greater, oral or equivalent, and cough and/or sore throat is used by CDC’s Outpatient Influenza-like Illness (ILI) Surveillance Network (ILINet), in which healthcare providers report the total number of patient visits and the number of patients seen for ILI each week.^9^

The DCM Precision Medicine Study, conducted by a multi-center consortium that is geographically dispersed across the United States, collected genetic and environmental information relevant to DCM. The study provided a unique opportunity to describe frequencies of DCM patients who experienced symptoms of flu-like illness prior to their diagnosis by demographics and clinical characteristics, and to explore if an antecedent flu-like illness modified the effect of genetics on DCM risk. We hypothesized that the relative risk of DCM in those who carried pathogenic, likely pathogenic, or variants of uncertain significance (P/LP/VUS) in DCM genes would be greater among those with an antecedent flu-like illness than among those without. We also hypothesized that the effects of common variants on DCM risk would be modified by having an antecedent flu-like illness.

## METHODS

### Data Availability

The data that support the findings of this study are available from the corresponding author upon reasonable request.

### The DCM Precision Medicine Study

The Precision Medicine DCM Study aimed to test the general hypothesis that DCM has substantial genetic basis.^10–13^ More than 1,200 patients with DCM (probands) meeting rigorous diagnostic criteria^10,14^ for DCM and approximately 2,000 of their relatives were identified and enrolled by heart failure and cardiac transplant physicians and clinical research personnel at multiple sites of the DCM Consortium.^10–13^ Accrual of patients with DCM (probands) and family members occurred from June 7, 2016, to March 15, 2020 (probands), and April 1, 2021 (family members). All enrolled probands had the diagnosis of DCM prior to the year 2020 when the COVID-19 pandemic started.

The Institutional Review Boards at the Ohio State University and the initial 12 clinical sites approved the study;^10^ oversight was ceded to a single IRB at the University of Pennsylvania as clinical sites beyond 12 were added.^12^ All participants provided written informed consent. This analysis used the data from all eligible probands aged 18 years and older who did not withdraw informed consent after recruitment and provided response to a flu-like illness history question at their enrollment interview (N=1164). Detailed inclusion and exclusion data for this study analysis are presented in Figure 1.

**Figure 1.**
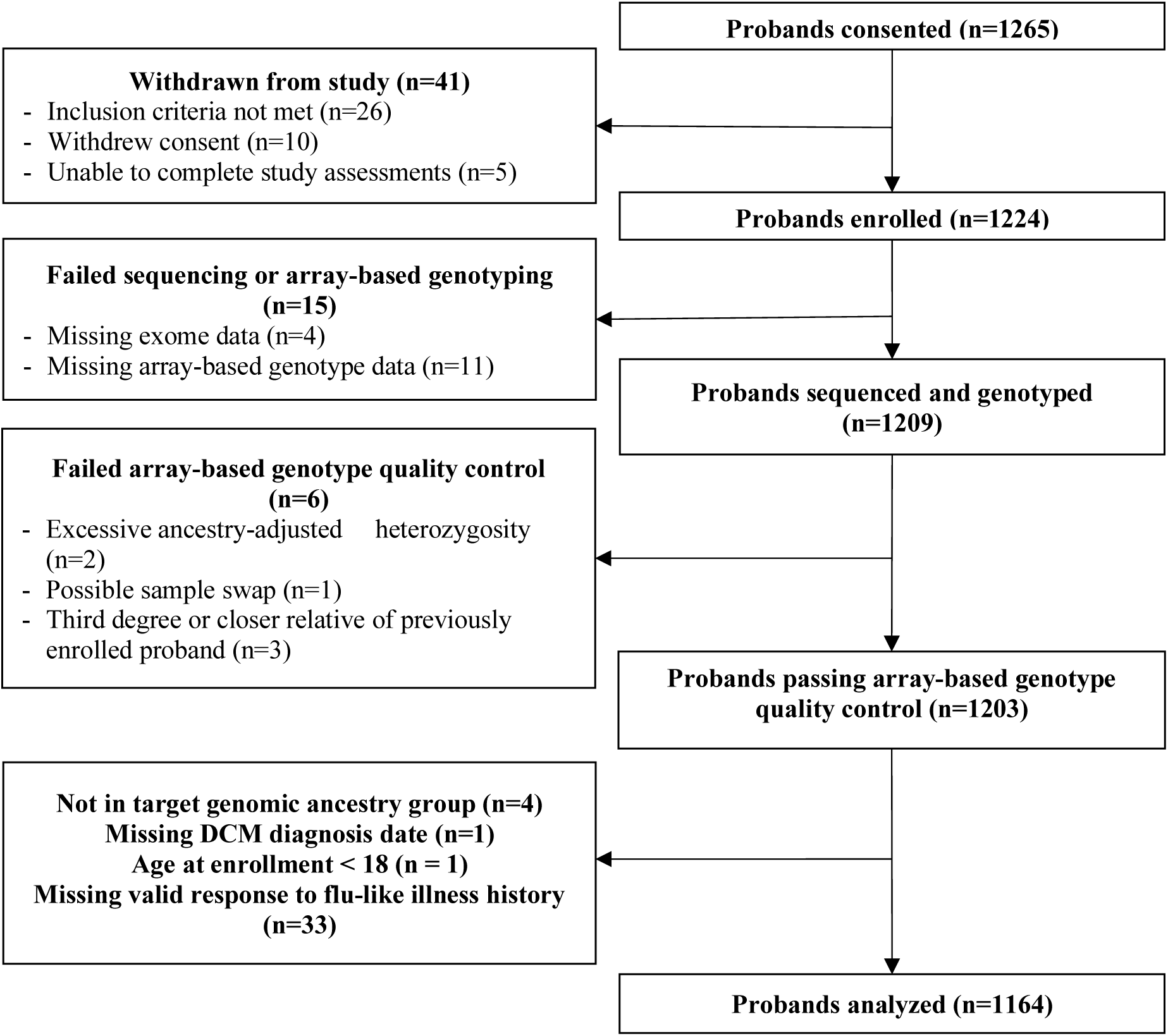
DCM Precision Medicine Study patients selected for the current analysis.

### Diagnostic criteria for DCM

All probands met diagnostic criteria for DCM, defined by a left ventricular ejection fraction <50% and a left ventricular internal diastolic dimension ≥95%th percentile for height and sex^15^, with other causes excluded, as previously defined.^10,12^ Cardiac magnetic resonance imaging data validated DCM diagnoses when available.^14^

### Medical history and demographic information

Structured interviews collected participants’ demographic and health history; medical record abstraction and review summarized cardiovascular clinical data.^10,12^ For patients with DCM, the date of diagnosis was obtained. Information on symptoms prior to the DCM diagnosis was obtained by a question, “*What* s*ymptoms did you have before you were diagnosed*?” A list of symptoms followed, and included *arrhythmia, presyncope, syncope, weight gain, edema, orthopnea, and dyspnea*. “*Episodes of flu-like illness (such as fever, sweats, chills, cough, achiness)*” was one of the listed symptoms.

The DCM Consortium is aware of recommendations for the collection, analysis, presentation and discussion of race, ethnicity, and ancestry and has adopted recommended approaches.^16^ Self-reported race and ethnicity data were obtained using structured race (Native American or Alaska Native, Asian, African American, Native Hawaiian or Pacific Islander, White, more than one race, or unknown) and Hispanic ethnicity (yes, no, or unknown) categories. This study presented both global genomic ancestry and self-reported race and ethnicity because of their relevance for health outcomes. Global genomic ancestry proportions were inferred from Illumina Global Screening Array genotypes using ADMIXTURE software^17^ with the 1000 Genomes Phase 3 integrated call set as the reference, as previously reported.^13^ An individual’s ancestry was defined as the continental ancestry group (African, European, Native American, or combined East and South Asian) accounting for the highest proportion of his or her genomic ancestry. Individuals with ancestry other than African, European, or Native American were not analyzed due to small numbers (n=4). Because genomic ancestry in these study participants is highly correlated with self-reported race and ethnicity (eTable 1), the results may be generalizable to clinical practice settings where self-reported information is the sole source for race and ethnicity definition.

### Genetic data collection

Research exome sequencing and analysis were conducted as described previously.^11,13^ Variants in 36 genes considered clinically relevant for DCM, including the 19 genes classified as definitive, strong, and moderate evidence by the Clinical Genome Resource (ClinGen),^18^ were assigned to an ACMG category: pathogenic (P), likely pathogenic (LP), variant of uncertain significance (VUS), likely benign, or benign; P, LP, and VUS variants were confirmed by Sanger sequencing.^11,13^ Genetic susceptibility in this analysis was measured by the presence of any DCM-related rare variants (P, LP, and VUS). In addition, analyses of more granular genetic risk groups based on the number of variant alleles or most deleterious variant harbored (P/LP, VUS only, and negative) were also conducted.

A genome-wide association study (GWAS) used Illumina Global Screening Array data from autosomal chromosomes for all probands. To improve statistical power and marker density, genotype data were imputed using the TOPMed Imputation Server with the TOPMed reference panel (version r2). Detailed descriptions of genotype quality control and imputation are provided in eAppendix 1. Ancestry principal component (PC) scores used to control for population stratification were obtained by projecting DCM Precision Medicine Study participants onto the PC space learned from the 1000 Genomes Phase 3 integrated callset, as previously reported.^13^

### Statistical analysis

All analyses were performed in R version 4.2.2 (R Foundation), Joinpoint software version 4.7.0.0., and SAS/STAT 15.2 software, Version 9.4 (TS1M7) of the SAS System for 64-bit Linux (SAS Institute). All statistical tests and p-values were two-sided with α=0.05 unless otherwise noted.

The percentages of DCM patients who had an antecedent flu-like illness prior to DCM diagnosis were calculated in groups defined by demographic characteristics (age at enrollment, gender, and race and ethnicity), ever use of tobacco, region of residence (Northeast, Midwest, West, South), and clinical characteristics such as major comorbidities and age at DCM diagnosis. To obtain model-based percentages in the presence of site heterogeneity, a logistic mixed model with a site random effect was fit using residual subject-specific pseudo-likelihood.^19,20^ Estimated percentages at a median US heart failure program (random effect of 0) and their 95% confidence intervals (CIs) were obtained using Morel-Bokossa-Neerchal bias-corrected empirical standard errors^19,21^ with sites as independent units and the standard normal distribution. Wald p-values and 95% CIs for subgroup percentage ratios were obtained using the delta method. To estimate means of LVIDD z-score and LVEF, a linear mixed model with a site random effect was fit using restricted maximum likelihood, and inference was performed using Morel-Bokossa-Neerchal bias-corrected empirical standard errors with sites as independent units and the standard normal distribution.

The association of the seasonal changes in percentages of antecedent flu-like illness with month of the self-reported DCM diagnosis was assessed by Joinpoint analysis.^22,23^ Of 1,164 patients enrolled, 735 (63.1%) patients provided a valid month of DCM diagnosis. Comparisons of these patients with those without reliable diagnosis months did not reveal significant differences in age at diagnosis or sex. The Joinpoint software (version 4.7.0.0) was used to fit weighted least-squares regression models to the monthly percentages of patients having a history of antecedent flu-like illness, with estimates of average monthly percent change (APC) and 95% CI.^22,23^ Using the default settings, which allowed for as few as two observed time points in the beginning, ending, and middle line segments (excluding the joinpoints), a maximum of 1 joinpoint was searched for using the Grid search algorithm and the permutation test and an overall alpha level of 0.05.

Because the flu-like illness question was only asked for those with a diagnosis of cardiomyopathy, a case-only design^24–26^ was used to examine whether antecedent flu-like illness modified the effect of genetic factors on DCM risk. A detailed description of this approach is provided in eAppendix 1. To summarize, case-only odds ratios (CORs) are estimated in probands under a logistic model with antecedent flu-like illness as the outcome and genetic risk factor as an explanatory variable with adjustment for site and potentially other covariates. If antecedent flu-like illness and genetic risk factors are not associated in the general population conditional on the adjustment variables, the COR estimates the interaction relative risk in the general population.^24–26^ Thus, if the COR is greater than (or less than) 1, the relative risk of DCM associated with a particular genetic risk factor would be higher (or lower) among those with an antecedent flu-like illness compared with those without. While such a design can identify whether an exposure modifies a genetic association with a phenotype when assumptions are met, it cannot estimate independent associations of either flu-like illness or genetics with DCM.^25,27^

To determine whether antecedent flu-like illness modified the effect of DCM-relevant rare variants on DCM risk, two logistic mixed models^19,20^ with antecedent flu-like illness as the outcome were fit to estimate CORs for each of the three genetic risk factor parameterizations (P/LP/VUS present or absent; P/LP, VUS only, or negative; counts of P/LP/VUS variant alleles). All models included a site random effect and were fit using residual subject-specific pseudo-likelihood; inference was performed using the Morel-Bokossa-Neerchal bias-corrected empirical covariance estimator^19,21^ with sites as independent units and the standard normal distribution. Model 1 estimated the crude COR; model 2 additionally adjusted for sex, predominant ancestry group (African, European, or Native American), age at diagnosis (<45, 45-64, ≥65), and tobacco use (ever, never).

To determine whether antecedent flu-like illness modified the association between each of 13,400,141 common/low-frequency autosomal variants (minor allele frequency ≥1%) and DCM, Firth logistic regression^28–30^ with a fixed effect for enrollment site was fit separately for each marker to estimate the COR in a case-only GWAS. The linear predictor also included the estimated alternate allele dosage of each marker, sex, age at diagnosis (<45, 45-64, ≥65), tobacco use (ever, never), the most deleterious rare variant harbored (LP/P, VUS, none), and 19 ancestry principal components as adjustment variables. Penalized profile likelihood 95% CIs and penalized likelihood ratio test p-values for the COR were obtained for each marker. A genome-wide significance level of 3×10^−8^ appropriate for lower-frequency common variants and trans-ethnic analyses was used.^31,32^ Additional GWAS details are provided in eAppendix 1.

## RESULTS

Of 1,164 adult DCM patients enrolled in the DCM Precision Medicine study, 515 (44.2%) were female and 480 (41.2%) were African American. The median time interval between DCM diagnosis and study enrollment was 5.3 years; 48.3% had the diagnosis less than 5 years ago. Overall, 351 (30.2%, 95% CI: 26.2% - 34.1%) reported experiencing symptoms of flu-like illness – such as fever, sweats, chills, cough, or achiness – prior to the diagnosis of cardiomyopathy (Table 1). At a median US advanced heart failure program, the estimated percentage of patients with a history of antecedent flu-like symptoms was higher among those aged <45 years at enrollment than those aged 45-64 year and those aged 65 years and older. The percentage also decreased by increasing age at diagnosis (P for trend = 0.009). Patients who had never smoked were less likely than those who had smoked to experience episodes of flu-like illness (28.2% vs. 32.4%, P=0.04). The percentages did not differ by sex and race and ethnicity. The percentages were also comparable with respect to selected major clinical characteristics such as hypertension, obesity, diabetes, cancer, and lung disease. The mean echo-based LVEF (%) at diagnosis was slightly lower (22.0 vs. 23.5, P=0.03) and LVIDD z-score was larger (4.5 vs 4.3, P=0.05) in patients who reported a flu-like illness prior to the DCM diagnosis compared with those without.

**Table 1.**
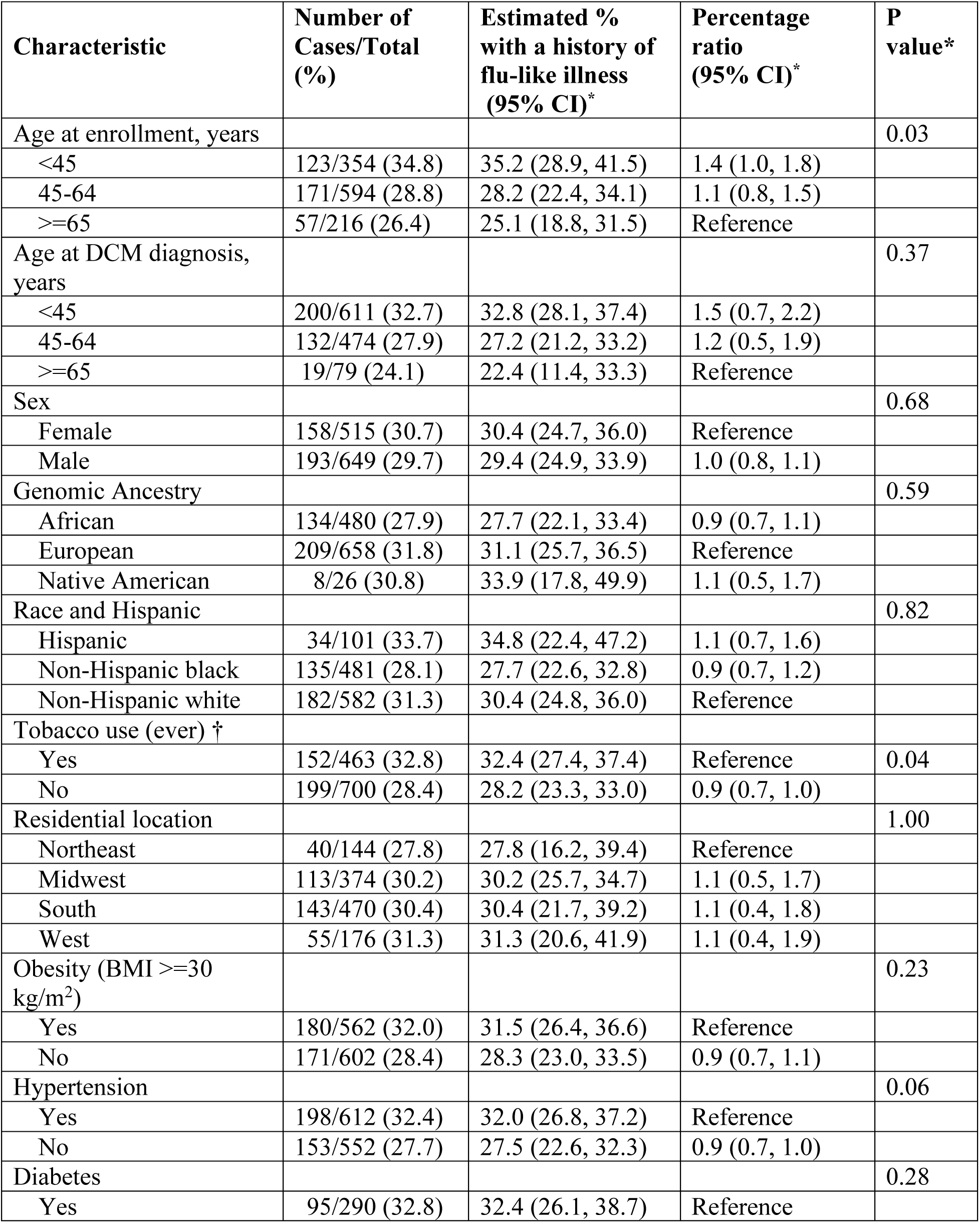
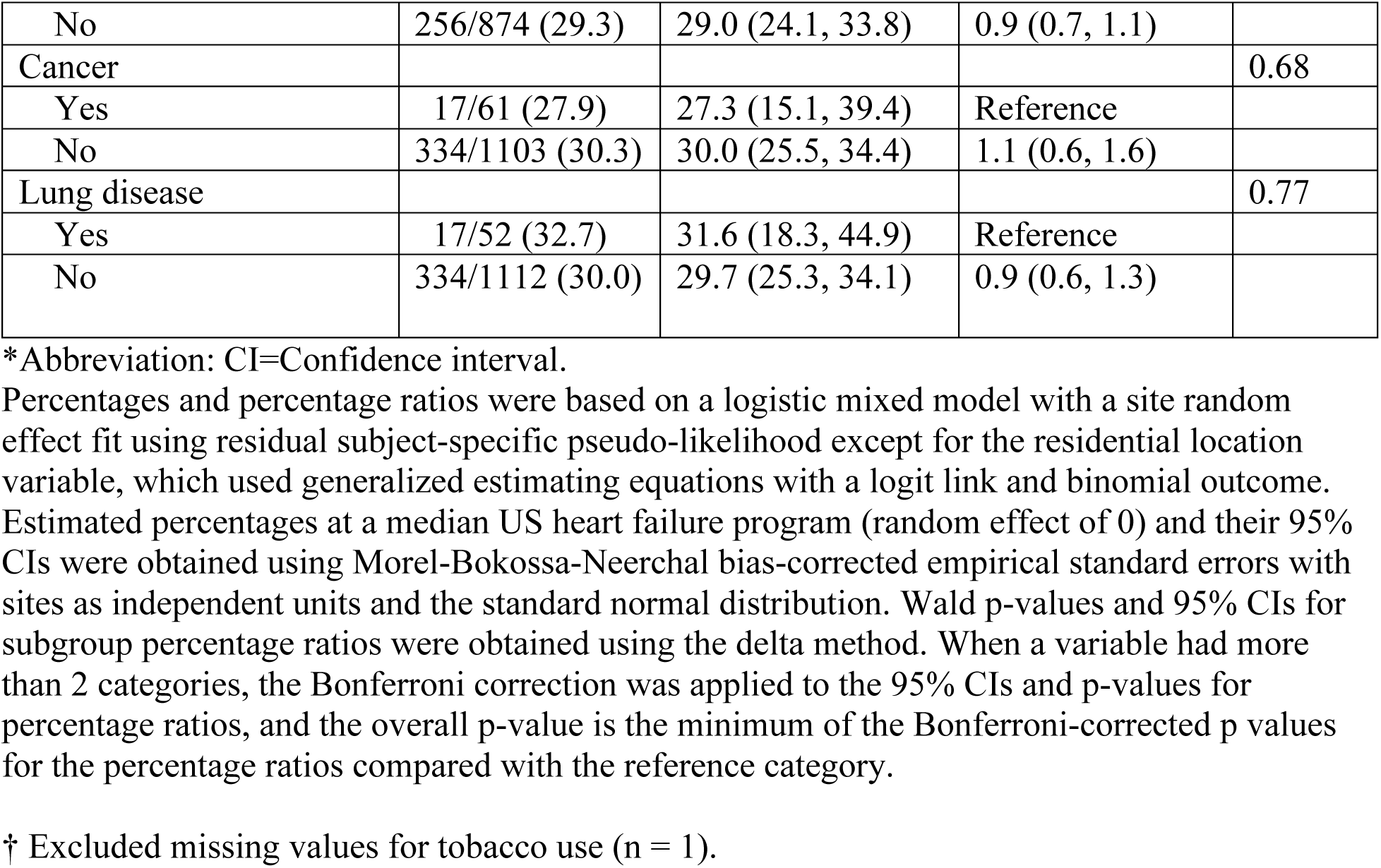
Percentage of patients with dilated cardiomyopathy who reported experiencing symptoms of flu-like illness prior to the diagnosis, by demographic and clinical characteristics.

| **Characteristic** | **Number of Cases/Total (%)** | **Estimated % with a history of flu-like illness (95% CI)\*** | **Percentage ratio (95% CI)\*** | **P value\*** |
| --- | --- | --- | --- | --- |
| Age at enrollment, years |  |  |  | 0.03 |
| <45 | 123/354 (34.8) | 35.2 (28.9, 41.5) | 1.4 (1.0, 1.8) |  |
| 45-64 | 171/594 (28.8) | 28.2 (22.4, 34.1) | 1.1 (0.8, 1.5) |  |
| >=65 | 57/216 (26.4) | 25.1 (18.8, 31.5) | Reference |  |
| Age at DCM diagnosis, years |  |  |  | 0.37 |
| <45 | 200/611 (32.7) | 32.8 (28.1, 37.4) | 1.5 (0.7, 2.2) |  |
| 45-64 | 132/474 (27.9) | 27.2 (21.2, 33.2) | 1.2 (0.5, 1.9) |  |
| >=65 | 19/79 (24.1) | 22.4 (11.4, 33.3) | Reference |  |
| Sex |  |  |  | 0.68 |
| Female | 158/515 (30.7) | 30.4 (24.7, 36.0) | Reference |  |
| Male | 193/649 (29.7) | 29.4 (24.9, 33.9) | 1.0 (0.8, 1.1) |  |
| Genomic Ancestry |  |  |  | 0.59 |
| African | 134/480 (27.9) | 27.7 (22.1, 33.4) | 0.9 (0.7, 1.1) |  |
| European | 209/658 (31.8) | 31.1 (25.7, 36.5) | Reference |  |
| Native American | 8/26 (30.8) | 33.9 (17.8, 49.9) | 1.1 (0.5, 1.7) |  |
| Race and Hispanic |  |  |  | 0.82 |
| Hispanic | 34/101 (33.7) | 34.8 (22.4, 47.2) | 1.1 (0.7, 1.6) |  |
| Non-Hispanic black | 135/481 (28.1) | 27.7 (22.6, 32.8) | 0.9 (0.7, 1.2) |  |
| Non-Hispanic white | 182/582 (31.3) | 30.4 (24.8, 36.0) | Reference |  |
| Tobacco use (ever) † |  |  |  |  |
| Yes | 152/463 (32.8) | 32.4 (27.4, 37.4) | Reference | 0.04 |
| No | 199/700 (28.4) | 28.2 (23.3, 33.0) | 0.9 (0.7, 1.0) |  |
| Residential location |  |  |  | 1.00 |
| Northeast | 40/144 (27.8) | 27.8 (16.2, 39.4) | Reference |  |
| Midwest | 113/374 (30.2) | 30.2 (25.7, 34.7) | 1.1 (0.5, 1.7) |  |
| South | 143/470 (30.4) | 30.4 (21.7, 39.2) | 1.1 (0.4, 1.8) |  |
| West | 55/176 (31.3) | 31.3 (20.6, 41.9) | 1.1 (0.4, 1.9) |  |
| Obesity (BMI >=30 kg/m<sup>2</sup>) |  |  |  | 0.23 |
| Yes | 180/562 (32.0) | 31.5 (26.4, 36.6) | Reference |  |
| No | 171/602 (28.4) | 28.3 (23.0, 33.5) | 0.9 (0.7, 1.1) |  |
| Hypertension |  |  |  | 0.06 |
| Yes | 198/612 (32.4) | 32.0 (26.8, 37.2) | Reference |  |
| No | 153/552 (27.7) | 27.5 (22.6, 32.3) | 0.9 (0.7, 1.0) |  |
| Diabetes |  |  |  | 0.28 |
| Yes | 95/290 (32.8) | 32.4 (26.1, 38.7) | Reference |  |
| No | 256/874 (29.3) | 29.0 (24.1, 33.8) | 0.9 (0.7, 1.1) |  |
| Cancer |  |  |  | 0.68 |
| Yes | 17/61 (27.9) | 27.3 (15.1, 39.4) | Reference |  |
| No | 334/1103 (30.3) | 30.0 (25.5, 34.4) | 1.1 (0.6, 1.6) |  |
| Lung disease |  |  |  | 0.77 |
| Yes | 17/52 (32.7) | 31.6 (18.3, 44.9) | Reference |  |
| No | 334/1112 (30.0) | 29.7 (25.3, 34.1) | 0.9 (0.6, 1.3) |  |

\*Abbreviation: CI=Confidence interval.
Percentages and percentage ratios were based on a logistic mixed model with a site random effect fit using residual subject-specific pseudo-likelihood except for the residential location variable, which used generalized estimating equations with a logit link and binomial outcome. Estimated percentages at a median US heart failure program (random effect of 0) and their 95% CIs were obtained using Morel-Bokossa-Neerchal bias-corrected empirical standard errors with sites as independent units and the standard normal distribution. Wald p-values and 95% CIs for subgroup percentage ratios were obtained using the delta method. When a variable had more than 2 categories, the Bonferroni correction was applied to the 95% CIs and p-values for percentage ratios, and the overall p-value is the minimum of the Bonferroni-corrected p values for the percentage ratios compared with the reference category.
† Excluded missing values for tobacco use (n = 1).

Figure 2 shows the number of DCM patients and the percentage who reported experiencing symptoms of flu-like illness prior to the diagnosis by month of their DCM diagnosis. The number of DCM occurrences fluctuated across the year, while the percentages of patients with episodes of flu-like illness tended to be lowest in August. However, the Joinpoint analysis did not reveal a trend of higher percentages in winter and fall months (APC for January-August = −3.96, P=0.1; APC for August to December =1.95, P=0.7). Figure 3 shows the percentage by region and age-at-diagnosis group. The percentages of patients who had symptoms of flu-like illness prior to the diagnosis were similar in the four geographic regions of residence among patients aged <45 years and 45-64 years. Percentages, however, differed for age 65 years and older with a higher rate in the South and West, although the differences did not reach statistical significance possibly due to small sample sizes.

**Figure 2.**
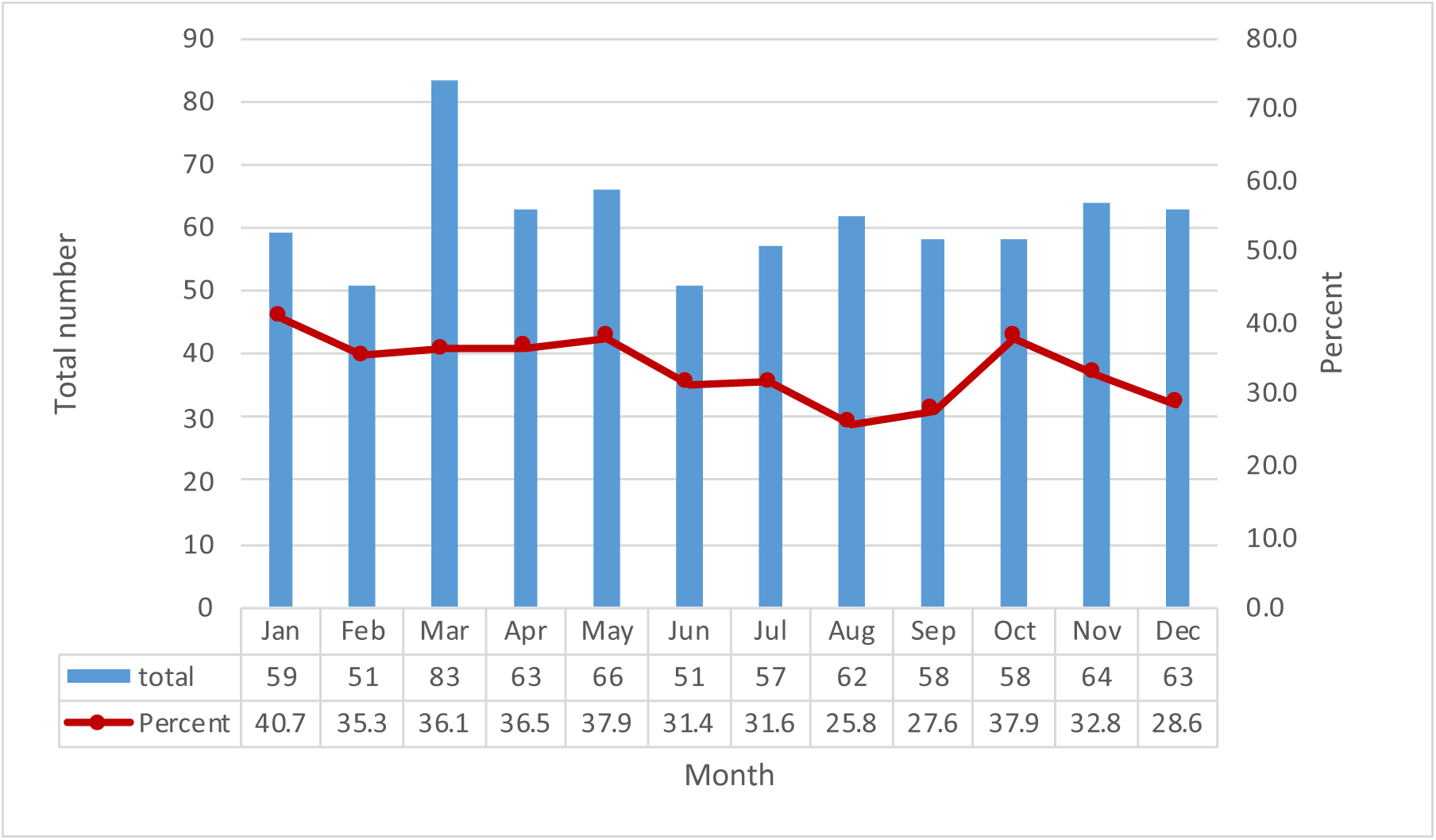
Number and percentage of patients with dilated cardiomyopathy who reported symptoms of flu-like illness prior to diagnosis by month of DCM diagnosis. Figure 2 shows the number of DCM patients and the percentage of them who experienced episodes of flu-like illness prior to the diagnosis by month of their DCM diagnosis. The number of DCM occurrences fluctuated across the year, while the percentages of patients who reported experiencing symptoms of flu-like illness tended to be lowest in August. The Joinpoint analysis did not reveal a trend of higher percentages in winter and fall months (APC for January-August = −3.96, P=0.1; APC for August to December =1.95, P=0.7). DCM=Dilated cardiomyopathy.

**Figure 3.**
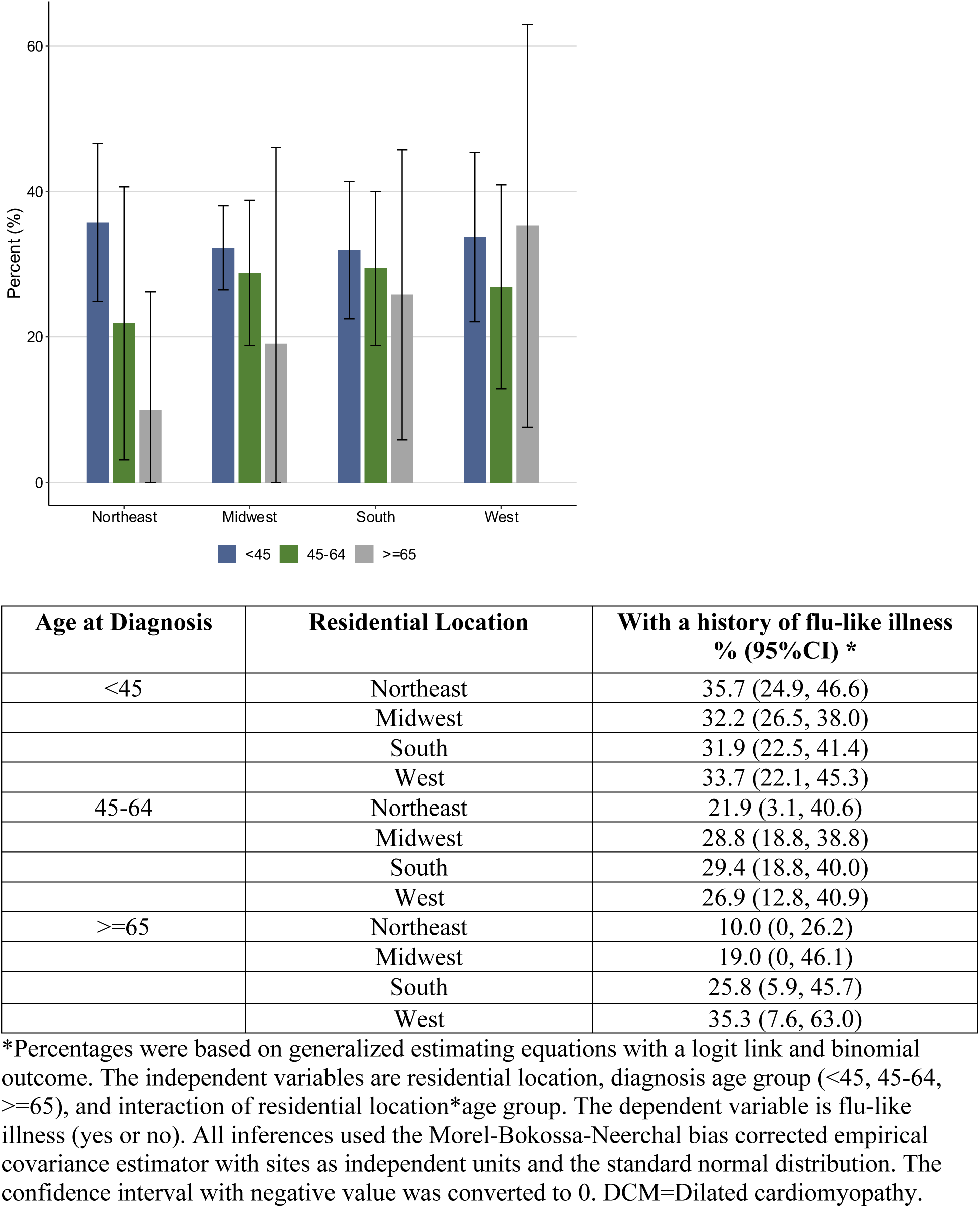
Percentage of patients with dilated cardiomyopathy who reported experiencing symptoms of flu-like illness prior to diagnosis by age at diagnosis and geographic location.

Table 2 presents percentages of patients who had symptoms of flu-like illness prior to the diagnosis by genetic risk. The percentage was 30.0% for those harboring a LP/P variant and 29.6% for those carrying VUS only, which were similar to those not harboring any P/LP/VUS (30.0%). Although the percentage (39.1%) was higher among those carrying 3 or more rare variants in DCM genes (P, LP or VUS) compared with those carrying none (30.0%), the difference was not statistically significant (percentage ratio 1.3, 95% CI=0.8-1.8). Frequencies of DCM patients harboring variants in specific DCM genes are provided in eTable 2 for those with and without a history of flu-like illness prior to the diagnosis.

**Table 2.**
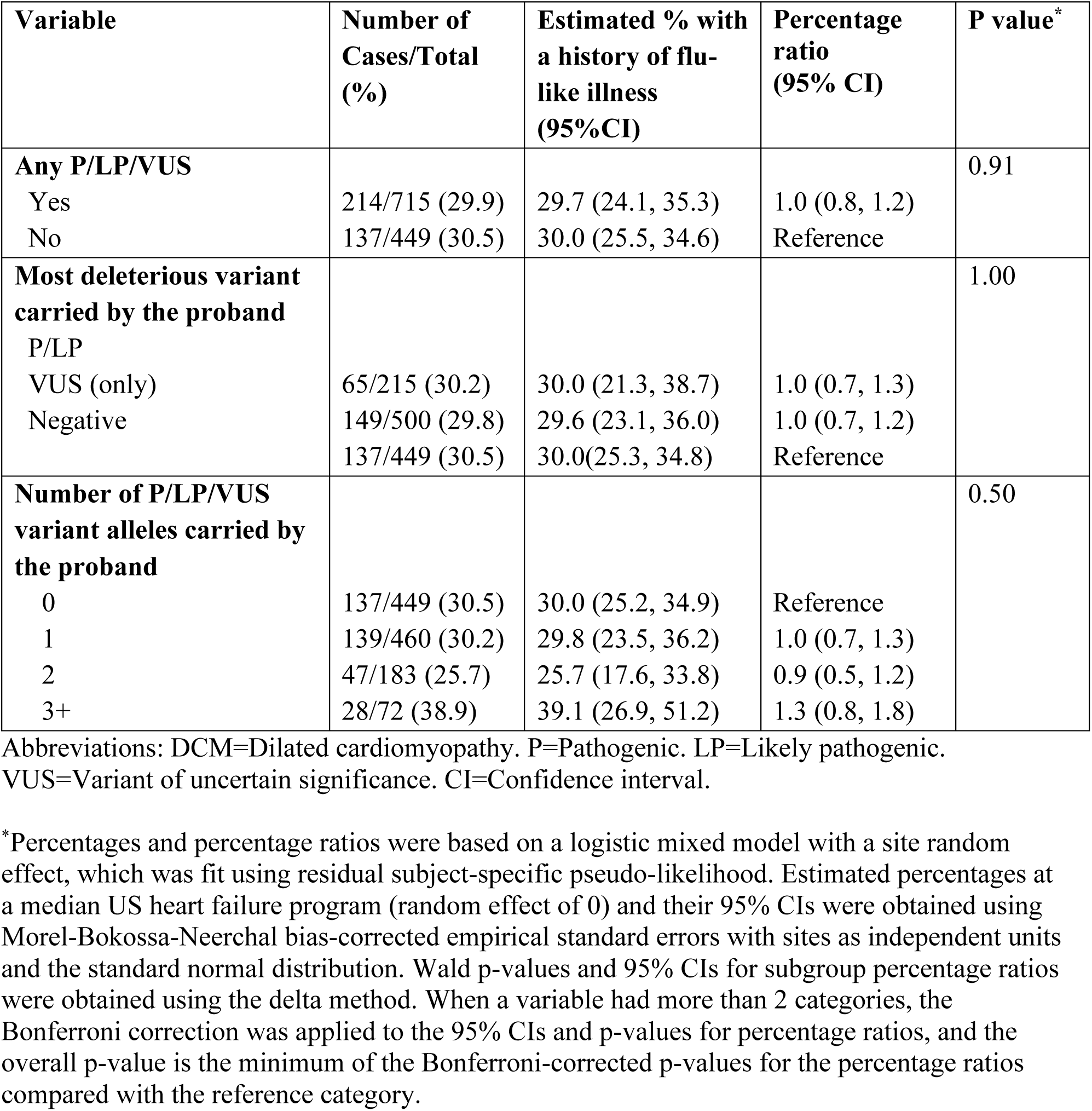
Percentages of patients with dilated cardiomyopathy who reported experiencing symptoms of flu-like illness prior to the diagnosis by presence of DCM-related rare variants.

| **Variable** | **Number of Cases/Total (%)** | **Estimated % with a history of flu-like illness (95%CI)** | **Percentage ratio (95% CI)** | **P value\*** |
| --- | --- | --- | --- | --- |
| **Any P/LP/VUS** |  |  |  | 0.91 |
| Yes | 214/715 (29.9) | 29.7 (24.1, 35.3) | 1.0 (0.8, 1.2) |  |
| No | 137/449 (30.5) | 30.0 (25.5, 34.6) | Reference |  |
| **Most deleterious variant carried by the proband** |  |  |  | 1.00 |
| P/LP |  |  |  |  |
| VUS (only) | 65/215 (30.2) | 30.0 (21.3, 38.7) | 1.0 (0.7, 1.3) |  |
| Negative | 149/500 (29.8) | 29.6 (23.1, 36.0) | 1.0 (0.7, 1.2) |  |
|  | 137/449 (30.5) | 30.0(25.3, 34.8) | Reference |  |
| **Number of P/LP/VUS variant alleles carried by the proband** |  |  |  | 0.50 |
| 0 | 137/449 (30.5) | 30.0 (25.2, 34.9) | Reference |  |
| 1 | 139/460 (30.2) | 29.8 (23.5, 36.2) | 1.0 (0.7, 1.3) |  |
| 2 | 47/183 (25.7) | 25.7 (17.6, 33.8) | 0.9 (0.5, 1.2) |  |
| 3+ | 28/72 (38.9) | 39.1 (26.9, 51.2) | 1.3 (0.8, 1.8) |  |

Abbreviations: DCM=Dilated cardiomyopathy. P=Pathogenic. LP=Likely pathogenic. VUS=Variant of uncertain significance. CI=Confidence interval.
\*Percentages and percentage ratios were based on a logistic mixed model with a site random effect, which was fit using residual subject-specific pseudo-likelihood. Estimated percentages at a median US heart failure program (random effect of 0) and their 95% CIs were obtained using Morel-Bokossa-Neerchal bias-corrected empirical standard errors with sites as independent units and the standard normal distribution. Wald p-values and 95% CIs for subgroup percentage ratios were obtained using the delta method. When a variable had more than 2 categories, the Bonferroni correction was applied to the 95% CIs and p-values for percentage ratios, and the overall p-value is the minimum of the Bonferroni-corrected p-values for the percentage ratios compared with the reference category.

Case-only odds ratios (CORs) are presented in Table 3. These estimate the interaction relative risk, which measures how much the relative risk for a given genetic risk level compared to the reference is higher (>1) or lower (<1) among those with antecedent flu-like illness than among those without. The crude COR for harboring any P/LP/VUS variant was consistent with the same relative risk in those with and without history of flu-like illness (1.0; 95% CI: 0.7-1.3). Adjusting for demographic variables including sex, predominant genomic ancestry, age at diagnosis, and ever use of tobacco did not alter this inference. Analyses considering the effects of the most deleterious variant or the number of P/LP/VUS alleles on DCM risk also did not reveal any statistically significant modifications of these effects (Table 3).

**Table 3.** Modification of the effects of DCM-related rare variants on the risk of dilated cardiomyopathy by antecedent flu-like illness.

| **Variables** | **Model 1\***<br>**COR (95% CI)** | **Model 2\***<br>**COR (95% CI)** |
| --- | --- | --- |
| **Any P/LP/VUS** |  |  |
| Yes | 1.0 (0.7, 1.3) | 0.9 (0.7, 1.3) |
| No | Reference | Reference |
| **Most deleterious variant carried by the proband** |  |  |
| P/LP | 1.0 (0.6, 1.5) | 0.9 (0.6, 1.5) |
| VUS (only) | 1.0 (0.7, 1.3) | 0.9 (0.7, 1.4) |
| Negative | Reference | Reference |
| **Number of P/LP/VUS variant alleles carried by the proband** |  |  |
| 0 | Reference | Reference |
| 1 | 1.0 (0.7, 1.4) | 0.9 (0.6, 1.4) |
| 2 | 0.8 (0.5, 1.2) | 0.8 (0.5, 1.3) |
| 3+ | 1.5 (0.9, 2.5) | 1.4 (0.8, 2.6) |

Abbreviations: DCM=Dilated cardiomyopathy. P=Pathogenic. LP=Likely pathogenic. VUS=Variant of uncertain significance. COR=Case-only odds ratio. CI=Confidence interval.
\*Estimates were obtained from logistic mixed models with a site random effect, which was fit using residual subject-specific pseudo-likelihood. The outcome is flu-like illness (yes or no) and the independent variables are different genetic risk parameterizations. All tests and 95% CIs used the Morel-Bokossa-Neerchal bias corrected empirical covariance estimator with sites as independent units and the standard normal distribution. The COR reflects the interaction relative risk, which measures how much the relative risk for that genetic risk level compared to the reference is higher ( $>1$ ) or lower ( $<1$ ) among those with antecedent flu-like illness than among those without. See eAppendix 1 for further details. Model 1 was unadjusted; model 2 adjusted for age group at diagnosis, sex, genomic ancestry and ever use of tobacco.

The case-only GWAS quantile-quantile plot (eFigure 1) did not show systematic inflation of type I error (genomic control λ=0.99), suggesting adequate control for population stratification. A genome-wide significant association was found at rs2102158 in chromosome 3q24 (P=2.74×10^−8^; Figure 4a, eTable 3); nearby genes are shown (Figure 4b). The next two strongest associations at rs60999026 (P=3.21×10^−7^) in 1p13.1 and rs2829363 (P=3.37×10^−7^) in 21q21.2 did not achieve genome-wide significance (eFigures 2 and 3; eTable 3).

**Figure 4.**
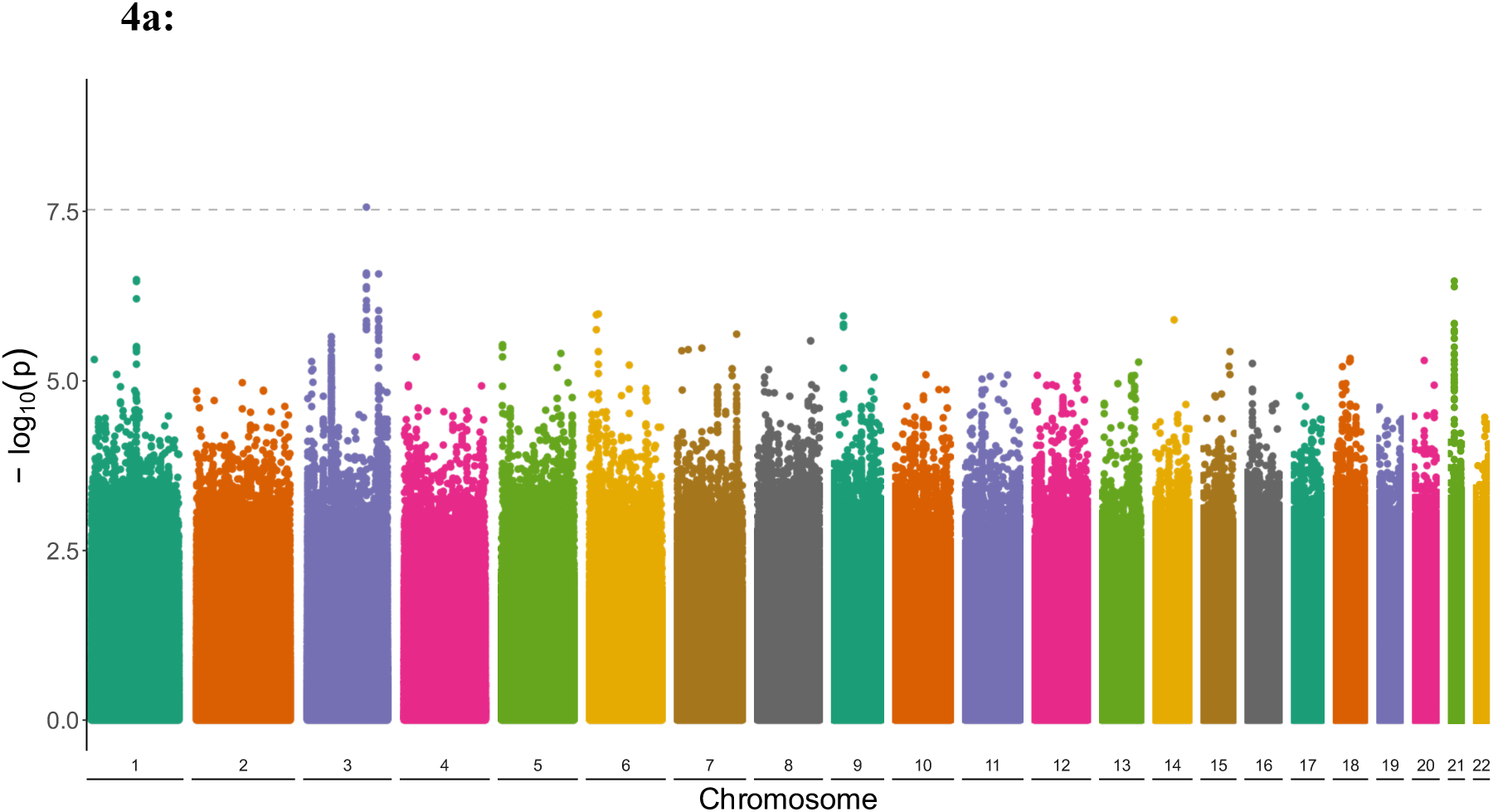
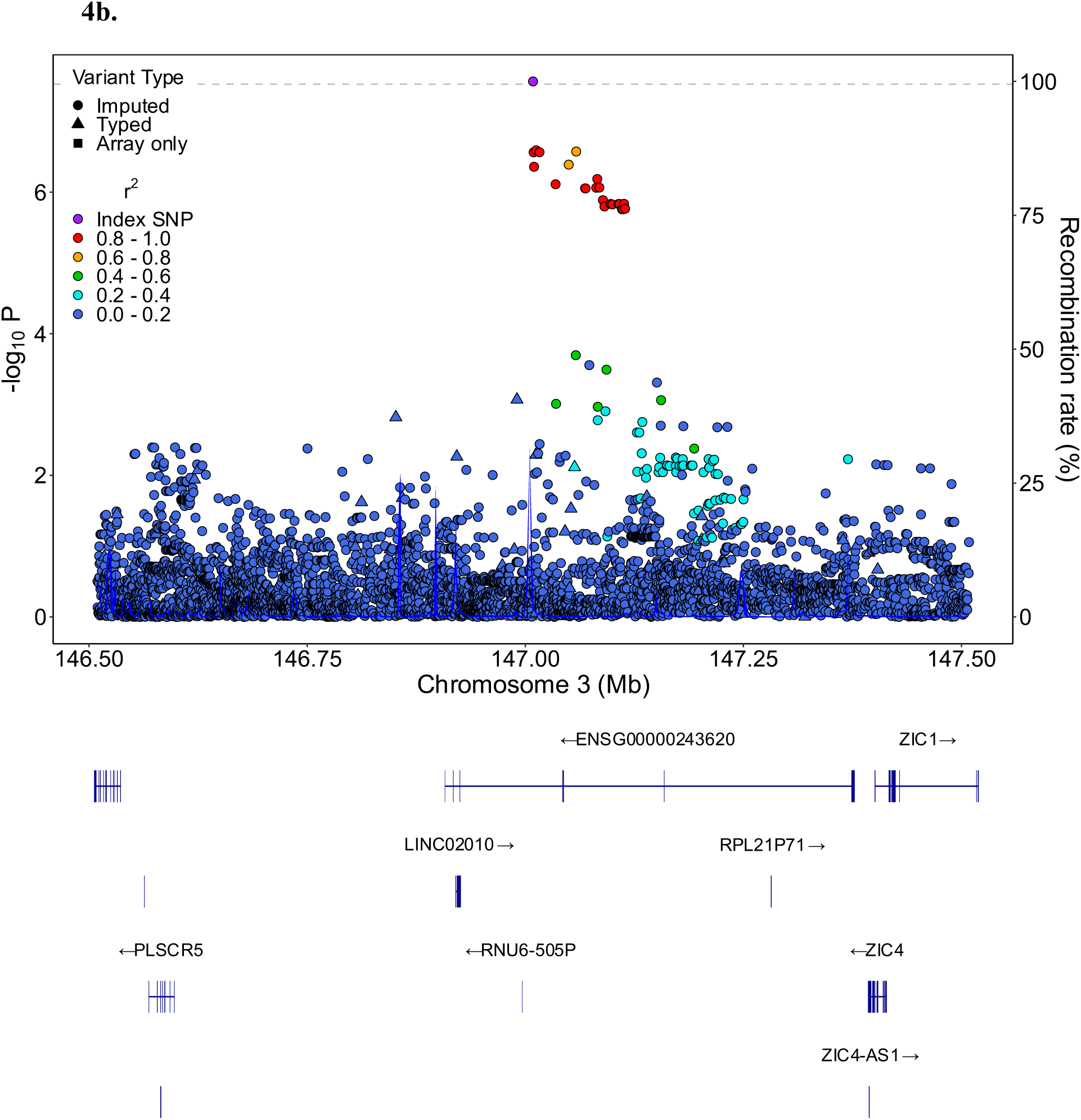
Modification of effects of common genetic variants on the risk of dilated cardiomyopathy by antecedent flu-like illness. **Figure 4a** provides a Manhattan plot of p-values for the case-only odds ratio for each marker. The peak on chromosome 3 reached genome-wide significance (3 × 10^−8^), indicated by the dashed line. Two other peaks found in chromosome 1 and chromosome 21 approached genome-wide significance (eTable 3; eFigures 2 and 3). **Figure 4b** shows the genome-wide significant peak in chromosome 3 in greater detail. The peak and the markers around the peak are from imputed variants.

## DISCUSSION

In this study we did not find evidence that episodes of flu-like illness prior to a DCM diagnosis modified the effect of harboring rare variants in DCM genes on DCM risk, even though approximately one third of patients with DCM reported symptoms of flu-like illness prior to the diagnosis. However, our GWAS analysis suggested that flu-like illness may modify the effect of a common variant on DCM risk. To our knowledge, this is the first study that provides a comprehensive epidemiologic profile of antecedent flu-like symptoms occurring among DCM patients prior to their diagnosis and explores whether flu-like illness modified genetic risk for those carrying DCM-related rare and common variants based on a large, geographically dispersed, and multi-race-ethnic consortium study.

Our results do not support the hypothesis that antecedent flu-like illness may be an effect modifier, or a trigger, that enhances the increase in DCM risk arising from DCM-relevant rare variants. In patients who carry pathogenic or likely pathogenic rare variants in DCM-relevant genes, the development of DCM occurs over time until DCM finally presents, in most cases with heart failure. DCM patients are known to have increased late gadolinium enhancement (LGE) found at cardiac magnetic resonance imaging, presumably related to ongoing inflammation from the genetic condition,^33^ with certain genetic backgrounds known to cause substantial LGE.^34^ One plausible mechanism of a flu-like illness triggering the individual’s transition from asymptomatic DCM to symptomatic heart failure could be added systemic disturbances such as fever and inflammation from influenza or a flu-like illness. This is well-supported by evidence that influenza may increase cardiovascular morbidity and mortality,^35^ with heart failure one of the most common complications of influenza.^35,36^ Our results indicate that a presentation of DCM following a flu-like illness does not signal a higher or lower rare variant genetic risk for DCM or alter the need for recommended genetic testing.

This finding is consistent with the study by Mahon et al who did not detect, using highly sensitive PCR procedures, myocardial viral infection among 19 relatives of patients with DCM who were suspected of having early disease.^4^ After comparing with 25 controls with ischemic heart disease, the authors concluded that there was no evidence that virus acted as a trigger factor for initiating DCM. This is also congruent with a more recent study to understand the role of viral infection in its association with cardiomyopathy.^37^

Nevertheless, prior evidence has suggested that influenza infection may play a causal role in the occurrence of incident or prevalent cardiovascular events, which is also supported by the cumulative evidence indicating the protective role of influenza vaccination against cardiovascular disease.^38,39^ Results from the recent COVID-19 pandemic also provided evidence that COVID-19 patients with one or more underlying conditions, including diabetes, hypertension, and cardiovascular disease, are more likely to become severely ill.^40^ While flu-like illness was not found to modify the effect of genetics on DCM, an infection itself could be a non-specific precipitant of DCM presentation with heart failure, as has recently been re-emphasized.^41^ Findings from this study do not address the long-held question of whether the clinical presentation of DCM can be caused by a flu-like illness in some patients; further investigation will be needed.

While the symptom recall for this study of flu-like symptoms might have represented viral etiology beyond influenza, this symptom complex is widely used epidemiologically to monitor influenza incidence.^8,9^ Influenza incidence is difficult to quantify precisely, as many or most of those infected may not seek medical attention and are therefore not formally diagnosed. An estimated annual influenza incidence of approximately 8% (varying from 3% to 11%) was derived by statistical extrapolation of US hospitalization data and a meta-analysis of published literature.^42^ In a recently conducted national online survey, 17.9% of adults aged 18-64 who received the flu vaccine reported flu or flu-like illness during the 2019-20 season.^43^ Our study revealed a remarkably higher percentage of DCM patients who reported experiencing symptoms of flu-like illness prior to the diagnosis compared with the estimates from population-based studies. This may reflect a tendency for healthcare providers to attribute nonspecific symptoms, such as dyspnea and cough, to an infection in younger people, rather than considering heart failure. The higher percentage with flu-like symptoms may also be attributable to differences in study populations in terms of age and health conditions in addition to differences in methodologies used to collect data and study time periods. For example, one study reported that 35% of patients with acute myocardial infarction had a history of preceding influenza or flu-like illness,^44^ which is slightly higher than the finding from the current study.

In this study, a case-only design was necessary because a history of flu-like symptoms was only obtained from participants (probands and family members) with a diagnosis of cardiomyopathy. Although we did find a higher percentage of patients with DCM who experienced symptoms of a flu-like illness prior to the diagnosis than would be expected based on population incidence, a case-only design does not permit assessment of the independent effect of flu-like illness on DCM risk. Instead, this study investigated whether antecedent flu-like illness modified the effect of genetics on DCM risk, a topic which has been controversial.^43,45–47^ These results from the large and diverse DCM Precision Medicine Study may help fill the gap in knowledge regarding the role of viral infection in DCM etiology.

The single genome-wide significant association suggested that the relative risk of DCM from each alternate allele of rs2102158 is greater among those with antecedent flu-like illness. While this could represent a genuine biological effect, it could also arise if rs2102158 is associated with occurrence of flu-like illness in the general population, which is plausible given the antiviral activity of the nearby gene *PLSCR1* in SARS-CoV-2^48^ and influenza^49^ infections. Replication of this genome-wide finding with alternative study designs would be needed to determine whether this constitutes a biological effect.

### Limitations

Several limitations should be considered in the interpretation of the study findings. First, the history of an antecedent flu-like illness in this study is based on patients’ self-report, which is a clinical case definition and without confirmatory diagnostic testing such as viral culture or polymerase chain reaction (PCR) for influenza. The infection rate of clinically-defined outcomes without laboratory confirmation of influenza could be difficult to interpret because of possible coincident occurrence of other respiratory pathogens (e.g., respiratory syncytial virus) or overlap symptoms with heart failure.^50^ In addition, we did not obtain a specific time course for the occurrence of flu-like illness symptoms prior to the DCM diagnosis. Further, about half of the patients had the DCM diagnosis more than 5 years ago. Because of the potential memory decay, their self-reported history of flu-like illness may have been under-estimated. However, the effect of this consideration on ascertaining the modifying effect of flu-like illness would be expected to be small because of the lack of evidence indicating the possibility of differential reporting bias by DCM genetic susceptibility.

Secondly, for the COR in this study design to provide a valid estimate of the interaction relative risk, there must be no association between DCM genetic susceptibility and flu-like viral infection in strata of the population defined by the adjustment variables.^26,27^ Although adjustment was made for important variables that may induce such associations, including genomic ancestry and clinical site, this assumption cannot be empirically tested. Additionally, the case-only design only allowed for the assessment of statistical interactions under a multiplicative model. Further studies that longitudinally collect the information on incident DCM and virus infection, or more general effects of any type of infection regardless of DCM status, may help address this scientific question.

The novel findings from the GWAS have not been replicated and are hypothesis - generating only, an essential step needed to validate the observations presented here. The lack of validation in large part is due to lack of a DCM study with genetics that also has clinical data regarding antecedent flu-like illness. Future DCM genetic studies should collect and expand on the information presented here.

### Conclusions

These limitations notwithstanding, this large study did not provide compelling evidence that antecedent flu-like illness modified the effect of genetics on the risk of DCM. Our findings warrant further investigation in epidemiological and functional studies to elucidate pathophysiology as well as to aid in the development of recommendations for DCM prevention.

## Data Availability

Data are available via dbGaP.

## Acknowledgements

The investigators thank the families with DCM who have participated in this study, without whom this effort would not be possible. The DCM Precision Medicine Study was supported by computational infrastructure provided by The Ohio State University Division of Human Genetics Data Management Platform and the Ohio Supercomputer Center. The case-only GWAS used the Trans-Omics in Precision Medicine (TOPMed) program imputation panel (version TOPMed-r2) supported by the National Heart, Lung and Blood Institute (NHLBI); see www.nhlbiwgs.org. TOPMed study investigators contributed data to the reference panel, which can be accessed through the TOPMed Imputation Server. The panel was constructed and implemented by the TOPMed Informatics Research Center at the University of Michigan (3R01HL-117626-02S1; contract HHSN268201800002I). The TOPMed Data Coordinating Center (3R01HL-120393-02S1; contract HHSN268201800001I) provided additional data management, sample identity checks, and overall program coordination and support. We gratefully acknowledge the studies and participants who provided biological samples and data for TOPMed.

## Sources of Funding

Research reported in this publication was supported by a parent award from the National Heart, Lung, And Blood Institute of the National Institutes of Health under Award Number R01HL128857 (Dr. Hershberger), which included a supplement from the National Human Genome Research Institute. The content is solely the responsibility of the authors and does not necessarily represent the official views of the National Institutes of Health. Support was also provided to the DCM Research Project from the Graciano Family Fund.

## Disclosures

None

## A List of the Supplemental Materials

Supplemental Methods

eTables 1-3

eFigures 1-3

Supplemental References

## Notes

### Competing Interest Statement

The authors have declared no competing interest.

### Author Declarations

The Institutional Review Boards at the Ohio State University and the initial 12 clinical sites approved the study;10 oversight was ceded to a single IRB at the University of Pennsylvania as clinical sites beyond 12 were added

### Summary of Updates

The manuscript has had minor changes and updates due to peer review. Also the Data Supplement has now been added.

