## Supplemental Material - Methods, Figures and Tables for "Antecedent flu-like illness and onset of idiopathic dilated cardiomyopathy: The DCM Precision Medicine Study"


**eAppendix 1.** Supplemental Methods

**eFigure 1.** QQ plot of p-values from case-only genome-wide association study

**eFigure 2**. Peak approaching genome-wide significance found at 1p13.1

**eFigure 3.** Peak approaching genome-wide significance found at 21q21.2

**eTable 1.** Consistency between genomic ancestry and self-reported race

**eTable 2.** Patients' genetic findings (P, LP, or VUS) by history of flu-like illness

**eTable 3.** Results from case-only genome-wide association study

**eAppendix 1. Supplemental Methods**

This work followed the STROBE guidelines for reporting observational studies.[1]

*Estimation of Interaction Relative Risk in a Case-Only Design*

A case-only analysis of probands can be used to estimate and test the interaction between a genetic risk factor and environmental risk factor on the relative risk scale.[2-4] The arguments of previous authors[2-4] are adapted below to allow for adjustment variables and site heterogeneity. For proband $j$ at site $k$, let $D_{jk}$ be an indicator variable taking the value of 1 if an individual has DCM and 0 otherwise, $Y_{jk}$ be an indicator variable taking the value of 1 if an individual reported antecedent flu-like illness and 0 otherwise, $G_{jk}$ denote alternate allele dosage or a vector of genetic risk category indicators, $Z_{jk}$ be a set of measured covariates that may influence DCM risk, and $U_k$ be unobserved site-level variables that may also influence DCM risk. If $Y_{jk} = 0$ and $G_{jk} = g_0$ are the reference values for antecedent flu-like illness and the genetic risk factor, the adjusted interaction relative risk, $IRR_g$, is the ratio of the relative risk of DCM with $G_{jk} = g$ compared to $G_{jk} = g_0$ among those with $Y_{jk} = 1$ to the same relative risk among those with $Y_{jk} = 0$. The interaction relative risk can be expressed as the following function of the conditional DCM probabilities:

$$IRR_g = \frac{\dfrac{\mathrm{P}(D_{jk} = 1 | Y_{jk} = 1, G_{jk} = g, Z_{jk}, U_k)}{\mathrm{P}(D_{jk} = 1 | Y_{jk} = 1, G_{jk} = g_0, Z_{jk}, U_k)}}{\dfrac{\mathrm{P}(D_{jk} = 1 | Y_{jk} = 0, G_{jk} = g, Z_{jk}, U_k)}{\mathrm{P}(D_{jk} = 1 | Y_{jk} = 0, G_{jk} = g_0, Z_{jk}, U_k)}} \quad (1)$$

Note that this is identical to the definition given by others, [2,4] $IRR_g = RR_{1g} / (RR_{0g} RR_{1g_0})$, where

$$RR_{yg} = \frac{\mathrm{P}(D_{jk} = 1 | Y_{jk} = y, g_{jk} = g, Z_{jk}, U_k)}{\mathrm{P}(D_{jk} = 1 | Y_{jk} = 0, g_{jk} = g_0, Z_{jk}, U_k)}$$

An application of Bayes' rule yields an alternative expression for the conditional probability of DCM:

$$\mathrm{P}(D_{jk} | Y_{jk}, G_{jk}, Z_{jk}, U_k) = \frac{\mathrm{P}(Y_{jk} | G_{jk}, Z_{jk}, U_k, D_{jk}) \mathrm{P}(D_{jk} | G_{jk}, Z_{jk}, U_k)}{\mathrm{P}(Y_{jk} | G_{jk}, Z_{jk}, U_k)}$$

Substitution into (1) and simplification yields:

$$IRR_g = \frac{\frac{\mathrm{P}\left(Y_{jk}=1\middle|G_{jk}=g,Z_{jk},U_k,D_{jk}=1\right)}{\mathrm{P}\left(Y_{jk}=1\middle|G_{jk}=g_0,Z_{jk},U_k,D_{jk}=1\right)}\frac{\mathrm{P}\left(D_{jk}=1\middle|G_{jk}=g,Z_{jk},U_k\right)}{\mathrm{P}\left(D_{jk}=1\middle|G_{jk}=g_0,Z_{jk},U_k\right)}}{\frac{\mathrm{P}\left(Y_{jk}=0\middle|G_{jk}=g,Z_{jk},U_k,D_{jk}=1\right)}{\mathrm{P}\left(Y_{jk}=0\middle|G_{jk}=g_0,Z_{jk},U_k,D_{jk}=1\right)}\frac{\mathrm{P}\left(D_{jk}=1\middle|G_{jk}=g,Z_{jk},U_k\right)}{\mathrm{P}\left(D_{jk}=1\middle|G_{jk}=g_0,Z_{jk},U_k\right)}} \times \frac{\frac{\mathrm{P}\left(Y_{jk}=1\middle|G_{jk}=g_0,Z_{jk},U_k\right)}{\mathrm{P}\left(Y_{jk}=1\middle|G_{jk}=g,Z_{jk},U_k\right)}}{\frac{\mathrm{P}\left(Y_{jk}=0\middle|G_{jk}=g_0,Z_{jk},U_k\right)}{\mathrm{P}\left(Y_{jk}=0\middle|G_{jk}=g,Z_{jk},U_k\right)}}$$
$$= \frac{\frac{\mathrm{P}\left(Y_{jk}=1\middle|G_{jk}=g,Z_{jk},U_k,D_{jk}=1\right)}{\mathrm{P}\left(Y_{jk}=0\middle|G_{jk}=g,Z_{jk},U_k,D_{jk}=1\right)}}{\frac{\mathrm{P}\left(Y_{jk}=1\middle|G_{jk}=g_0,Z_{jk},U_k,D_{jk}=1\right)}{\mathrm{P}\left(Y_{jk}=0\middle|G_{jk}=g_0,Z_{jk},U_k,D_{jk}=1\right)}} \frac{\frac{\mathrm{P}\left(Y_{jk}=1\middle|G_{jk}=g_0,Z_{jk},U_k\right)}{\mathrm{P}\left(Y_{jk}=0\middle|G_{jk}=g_0,Z_{jk},U_k\right)}}{\frac{\mathrm{P}\left(Y_{jk}=1\middle|G_{jk}=g,Z_{jk},U_k\right)}{\mathrm{P}\left(Y_{jk}=0\middle|G_{jk}=g,Z_{jk},U_k\right)}} \quad (2)$$

The first term on the righthand side of (2) is the adjusted odds ratio comparing the odds of $Y_{jk} = 1$ between cases with $G_{jk} = g$ and $G_{jk} = g_0$ within strata defined by $\{Z_{jk}, U_k\}$, which we refer to as the case-only odds ratio, $COR_g$. The second term in (2) is an odds ratio describing the association between $Y_{jk}$ and $G_{jk}$ in the general population within strata defined by $\{Z_{jk}, U_k\}$. Upon further assuming that $Y_{jk}$ and $G_{jk}$ are conditionally independent within strata defined by $\{Z_{jk}, U_k\}$ in the population, $\mathrm{P}(Y_{jk}|G_{jk}, Z_{jk}, U_k) = \mathrm{P}(Y_{jk}|Z_{jk}, U_k)$, and this odds ratio in (2) equals 1, leaving:

$$IRR_g = \frac{\mathrm{P}\left(Y_{jk}=1\middle|G_{jk}=g,Z_{jk},U_k,D_{jk}=1\right)}{\mathrm{P}\left(Y_{jk}=0\middle|G_{jk}=g,Z_{jk},U_k,D_{jk}=1\right)} \Bigg/ \frac{\mathrm{P}\left(Y_{jk}=1\middle|G_{jk}=g_0,Z_{jk},U_k,D_{jk}=1\right)}{\mathrm{P}\left(Y_{jk}=0\middle|G_{jk}=g_0,Z_{jk},U_k,D_{jk}=1\right)} \quad (3)$$

Equation (3) implies that $IRR_g = COR_g$, which can be estimated with only probands using a logistic model for the odds of flu-like illness.

To specify a logistic model for $\mathrm{P}(Y_{jk} = 1|G_{jk}, Z_{jk}, U_k, D_{jk} = 1)$, we included vectors of fixed effects for $G_{jk}$ and $Z_{jk}$ as well as a site-specific intercept capturing the effect of the unobserved $U_k$:

$$\log\left(\frac{\mathrm{P}\left(Y_{jk}=1\middle|G_{jk},Z_{jk},U_k,D_{jk}=1\right)}{\mathrm{P}\left(Y_{jk}=0\middle|G_{jk},Z_{jk},U_k,D_{jk}=1\right)}\right) = \alpha_k + G_{jk}\beta + Z_{jk}\gamma \quad (4)$$

where $\beta$ and $\gamma$ are possibly vector-valued fixed effects and $\alpha_k$ is a site-specific intercept reflecting the effect of $U_k$.

The site-specific intercepts $\alpha_k$ can be modeled as independent $N(0, \sigma^2_{\text{site}})$ random effects in a generalized linear mixed model (GLMM) framework,[5,6] which allows generalization to the larger population of US advanced heart failure programs. The random effect approach assumes that the site-specific intercepts $\alpha_k$ induced by the $U_k$ are independent of $G_{jk}$ and $Z_{jk}$ among probands.[7-11] However, even if the $U_k$ are independent of $G_{jk}$, $Z_{jk}$, and $Y_{jk}$ in the population, they might not be independent of them among subjects with $D_{jk} = 1$ due to conditioning on a common effect.[12] While this is a concern in setting with a small number of observations per independent unit, violation of this assumption is unlikely to induce substantial bias in estimates of $\beta$ and $\gamma$ with moderate or large numbers of probands per site.[9-11] Because the current study had a relatively large number of probands per site (mean 41; median 31; IQR: 19 – 49), GLMM was a viable approach that was used for analyses examining modification of the effects of rare variants by antecedent flu-like illness, as described in the main text.

Unfortunately, the GLMM approach does not scale well to a genome-wide association study (GWAS) because finite-sample estimation issues such as separation and non-convergence can arise frequently when fitting a GLMM for each of millions of variants. Under the asymptotic model above, the site-specific intercepts $\alpha_k$ could instead be modeled as fixed effects using Firth penalized maximum likelihood (FPML).[8,13] While this approach limits inferences to the sites involved in this study, it does not assume that the site-specific intercepts $\alpha_k$ induced by the $U_k$ are independent of $G_{jk}$ and $Z_{jk}$ and also has additional advantages in the GWAS context. The primary advantage is that the Firth correction applies to all coefficients, thereby automatically resolving finite-sample issues such as infinite $\alpha_k$ for sites with $Y_{jk} = 0$ or $Y_{jk} = 1$ for all $j$[8] and separation due to low-frequency variants[14] as part of estimation.[15,16] A secondary advantage is computational feasibility,[14] including the ability to use the established implementation from the logistf R package within the GWASTools Bioconductor package.[17] FPML was therefore used to estimate the model in (4) with fixed site effects for the case-only GWAS, as described in further detail below.

*Genotyping and Imputation*

Illumina Global Screening Array genotyping and quality control (QC) procedures for these samples have been described in detail previously.[18] Briefly, these procedures included filters on marker and sample call rate, marker and family Mendelian inconsistencies, sample ancestry-adjusted autosomal heterozygosity, sample sex discordance, and sample relatedness compared with pedigree expectations. High-quality biallelic autosomal variants passing these QC steps with call rates of at least 90% among 1524 subjects passing QC were selected for imputation. Before imputation, monomorphic variants and indels were removed, and triple-liftOver (version 1.33) was used to convert genome coordinates from GRCh37 to GRCh38 with a procedure that correctly handles variants in regions inverted between the two reference genome builds.[19] Pre-imputation filtering of variants on Hardy-Weinberg Equilibrium (HWE) has been shown to have minimal effect on imputation quality[20,21] and was therefore not performed, especially since a deviation from HWE in a case-only sample can occur for variants associated with disease under a non-multiplicative model.[22,23] The TOPMed Imputation Server (pipeline version 1.7.4) with the TOPMed reference panel (version r2) was used to perform additional quality control, pre-phasing, and imputation.[24-26] Some variants were removed before or during the phasing step due to decomposed multi-allelic variants or not being included in the TOPMed reference panel used for phasing. Because genotypes for these variants were either fully removed from the imputed data set or fully imputed without using any of the original genotypes, any imputed genotypes were discarded, and the original genotypes were added back to the VCF post-imputation. The resulting final merged set of 292,143,416 variants included 492,078 typed and imputed, 291,623,690 fully imputed, and 27,648 with array genotypes only.

*Case-Only Genome-wide Association Study*

A case-only genome-wide association study (GWAS) was performed in 1163 probands with complete covariate data to assess whether a history of flu-like illness at the time of DCM diagnosis modified the effect of common/low-frequency variants on the odds of DCM.[27,28] Fully imputed variants with imputation $R^2 \geq 0.3$, typed and imputed variants with empirical $R^2 \geq 0.9$ and imputation $R^2 \geq 0.3$, and variants with only array genotypes, all with sample minor allele frequency (MAF) ≥1% calculated from the estimated alternate allele dosage (DS), were selected. Variants were not further filtered based on deviations from HWE because these can occur for disease-associated variants within cases, as noted above.[22,23] The final set of 13,400,141 variants was 54.6% common (MAF ≥5%) and 45.4% low-frequency (MAF within 1-5%) with an average imputation $R^2$ of 0.96 among fully imputed variants and typed and imputed variants, and an average empirical $R^2$ of 0.97 among typed and imputed variants.

Firth logistic regression[13-15] was fit for each variant with the indicator for a history of flu-like illness at the time of DCM diagnosis as the response and estimated alternate allele dosage, sex, age at diagnosis (<45, 45-64, ≥65), tobacco use (ever, never), most deleterious rare variant harbored (LP/P, VUS only, none), 19 ancestry principal components (PCs), and enrollment site as fixed effects. Ancestry PCs were determined by projecting probands onto the 1000 Genomes PC space, as described previously.[18] Because fewer than 20 PCs could have been sufficient to describe variation in this sample with only a subset of 1000 Genomes ancestries, non-redundant PCs were determined by sequentially linearly regressing $PC_k$ (starting with $PC_2$ because $PC_1$ was always considered non-redundant) on all PCs up to $k - 1$ with an intercept. If $R^2 \geq 0.95$, then the PC was considered redundant and eliminated from subsequent regressions, otherwise the PC was kept. All models were fit using the logistf R package (version 1.26.0) within the GWASTools Bioconductor package (version 1.44.0).[17] Penalized profile likelihood 95% confidence intervals and penalized likelihood ratio p-values for the COR were obtained for each marker.[29] A lower genome-wide significance threshold of $3 \times 10^{-8}$ more appropriate for lower-frequency common variants and trans-ethnic analyses was used;[30,31] this was more conservative than the standard genome-wide significance threshold of $5 \times 10^{-8}$.[30-33]

LocusZoom plots were made using locuszoomr R package (version 0.3.0) to show variants around the peaks with annotation based on Ensembl version 105. The recombination rate was included from "recomb1000GAvg" in Recombination Rate Tracks page of UCSC.

**eFigure 1. QQ plot of p-values from case-only genome-wide association study**

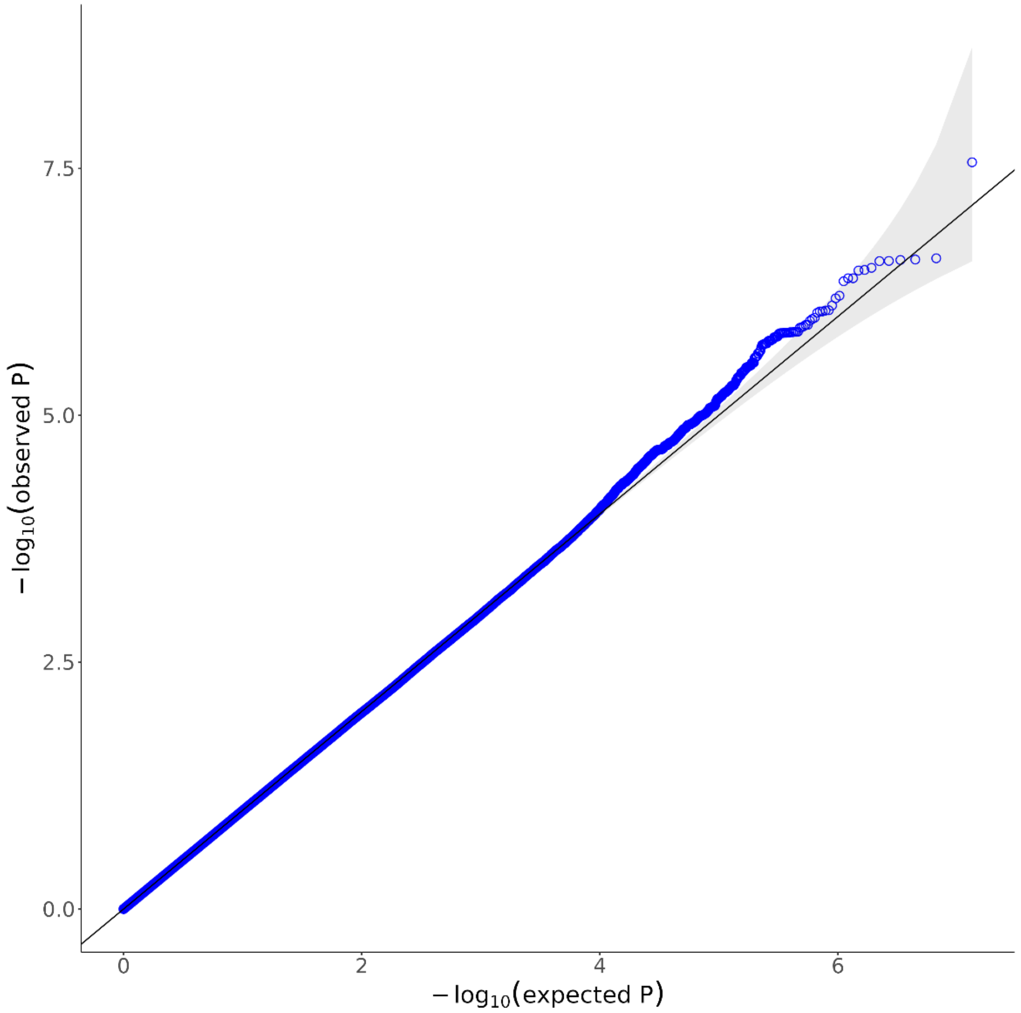

**eFigure 2. Peak approaching genome-wide significance at 1p13.1**

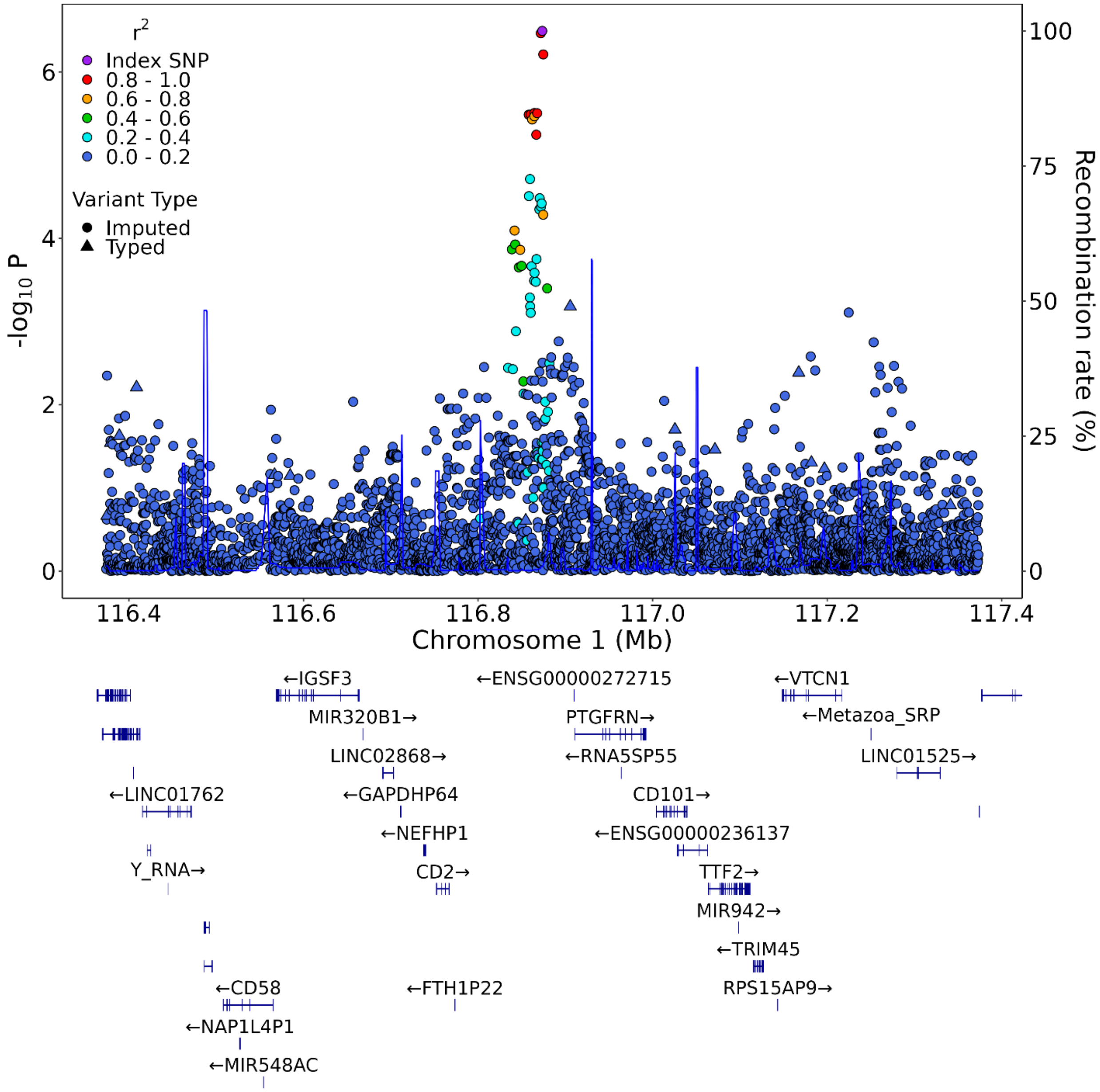

**eFigure 3. Peak approaching genome-wide significance at 21q21.2**

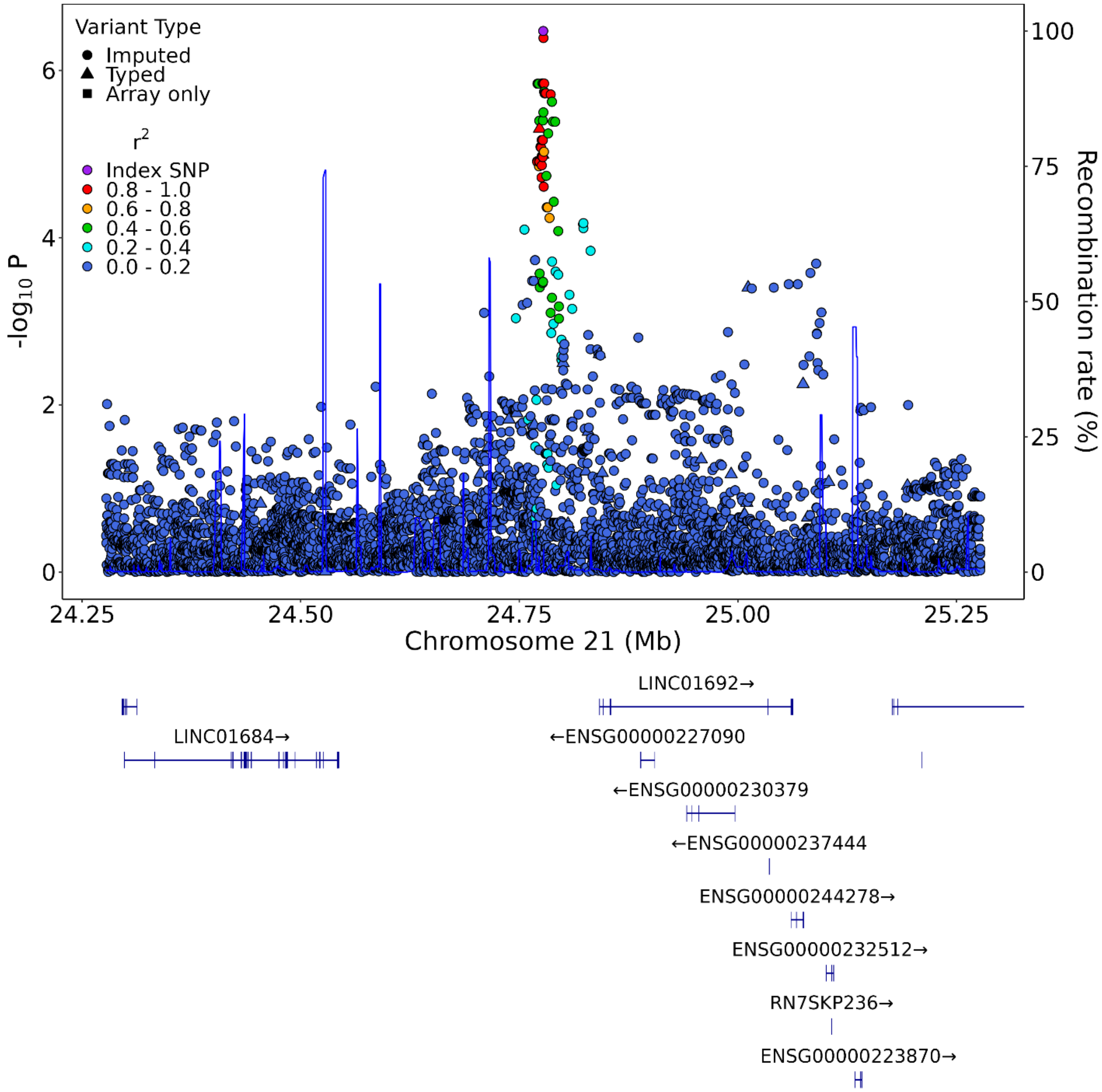

**eTable 1. Consistency between genomic ancestry and self-reported race**

| Genomic ancestry | Self-reported race | | | |
|---|---|---|---|---|
| | African American<br>N (%) | White<br>N (%) | More than one race<br>N (%) | Total<br>N (%) |
| **Total** | 490 (100.0) | 672 (100.0) | 2 (100.0) | 1164 (100.0) |
| **African** | 479 (97.8) | 1 (0.1) | 0 (0.0) | 480 (41.2) |
| **European** | 10 (2.0) | 647 (96.3) | 1 (50.0) | 658 (56.5) |
| **Native American** | 1 (0.2) | 24 (3.6) | 1 (50.0) | 26 (2.2) |

**eTable 2. Patients' genetic findings (P, LP, or VUS) by history of antecedent flu-like illness**

| Gene | With flu-like illness history N = 351 | Without flu-like illness history N = 813 |
|---|---|---|
| *ABCC9* | 4 (1.1) | 10 (1.2) |
| *ACTC1* | 0 (0) | 1 (0.1) |
| *ACTN2* | 4 (1.1) | 12 (1.5) |
| *ANKRD1* | 6 (1.7) | 8 (1.0) |
| *BAG3* | 6 (1.7) | 16 (2.0) |
| *CRYAB* | 0 (0) | 0 (0) |
| *CSRP3* | 0 (0) | 5 (0.6) |
| *DES* | 1 (0.3) | 8 (1.0) |
| *DSG2* | 8 (2.3) | 24 (3.0) |
| *DSP* | 24 (6.8) | 58 (7.1) |
| *EYA4* | 3 (0.9) | 8 (1.0) |
| *FLNC* | 34 (9.7) | 53 (6.5) |
| *ILK* | 3 (0.9) | 2 (0.2) |
| *JPH2* | 3 (0.9) | 7 (0.9) |
| *LAMA4* | 9 (2.6) | 29 (3.6) |
| *LDB3* | 9 (2.6) | 25 (3.1) |
| *LMNA* | 10 (2.8) | 27 (3.3) |
| *MYBPC3* | 18 (5.1) | 39 (4.8) |
| *MYH6* | 19 (5.4) | 42 (5.2) |
| *MYH7* | 16 (4.6) | 36 (4.4) |
| *MYPN* | 10 (2.8) | 15 (1.8) |
| *NEBL* | 7 (2.0) | 15 (1.8) |
| *NEXN* | 2 (0.6) | 12 (1.5) |
| *PDLIM3* | 3 (0.9) | 5 (0.6) |
| *PKP2* | 11 (3.1) | 18 (2.2) |
| *PLN* | 0 (0) | 1 (0.1) |
| *RBM20* | 15 (4.3) | 31 (3.8) |
| *SCN5A* | 13 (3.7) | 38 (4.7) |
| *SGCD* | 3 (0.9) | 2 (0.2) |
| *TCAP* | 1 (0.3) | 11 (1.4) |
| *TNNC1* | 1 (0.3) | 4 (0.5) |
| *TNNI3* | 3 (0.9) | 5 (0.6) |
| *TNNT2* | 6 (1.7) | 13 (1.6) |
| *TPM1* | 2 (0.6) | 6 (0.7) |
| *TTN* | 62 (17.7) | 115 (14.1) |
| *VCL* | 5 (1.4) | 13 (1.6) |

NOTE: Each cell represents the number of individuals within each group (probands with and without history of flu-like illness) harboring at least one P/LP/VUS variant in a particular gene. The sum of column percentages may exceed 1 because an individual can harbor variants in more than one gene.

**eTable 3. Results from case-only genome-wide association study**

| Top Hits (GRCh38) | Variant identifiers | Chromosome Band | REF Allele | ALT Allele | Case-only odds ratio per alternate allele, 95% CI | P value |
|---|---|---|---|---|---|---|
| chr1:116873559:T:C | rs60999026 | 1p13.1 | T | C | 2.22 (1.63, 3.05) | $3.21x10^{-7}$ |
| chr3:147008842:G:T | rs2102158 | 3q24 | G | T | 2.72 (1.92, 3.89) | $2.74x10^{-8}$ |
| chr21:24777383:T:C | rs2829363 | 21q21.2 | T | C | 1.94 (1.51, 2.51) | $3.37x10^{-7}$ |
